# Predicting transitions between longitudinal classes of posttraumatic stress disorder, adjustment disorder, and well-being during the COVID-19 pandemic: Protocol of a latent transition model in a general Dutch sample

**DOI:** 10.1101/2021.08.30.21262819

**Authors:** Lonneke I.M. Lenferink, Joanne Mouthaan, Anna M. Fritz, Suzan Soydas, Marloes Eidhof, Marie-José van Hoof, Simon Groen, Trudy Mooren

**Affiliations:** Department of Psychology, Health, & Technology, Faculty of Behavioural, Management, and Social Sciences, University of Twente, Drienerlolaan 5, 7522 NB Enschede, the Netherlands; Department of Clinical Psychology and Experimental Psychopathology, Faculty of Behavioral and Social Sciences, University of Groningen, Grote Kruisstraat 2/1, 9712 TS Groningen, the Netherlands; Department of Clinical Psychology, Faculty of Social Sciences, Utrecht University, P.O. Box 80140, 3508 TC, Utrecht, the Netherlands; Department of Clinical Psychology, Institute of Psychology, Faculty of Social Sciences, Leiden University, Leiden, The Netherlands; ARQ Centre ‘45, partner in ARQ National Psychotrauma Centre, Diemen, the Netherlands; Reinier van Arkel Psychotraumacenter South Netherlands, Den Bosch, The Netherlands; Radboud University, Behavioural Science Institute, Nijmegen, The Netherlands; Amsterdam UMC, Department of Child and Adolescent Psychiatry, Meibergdreef, Amsterdam, The Netherlands; LUMC-Curium, Department of Child and Adolescent Psychiatry, Albinusdreef 2, Leiden, The Netherlands; Centre for Transcultural Psychiatry, GGZ Drenthe Mental Health Care, De Evenaar, Centre for Transcultural Psychiatry, Beilen, the Netherlands

**Keywords:** COVID-19, stressors, pandemic, adjustment disorder, post-traumatic stress disorder, well-being, predictors, trajectory, transition

## Abstract

**Background:** A growing body of literature shows profound effects of the COVID-19 pandemic on mental health, among which increased rates of posttraumatic stress disorder (PTSD) and adjustment disorder (AD). However, current research efforts have largely been unilateral, focusing on psychopathology and not including well-being, and are dominated by examining average psychopathology levels or on disorder absence/presence, thereby ignoring individual differences in mental health. Knowledge on individual differences, as depicted by latent subgroups, in the full spectrum of mental health may provide valuable insights in how individuals transition between health states and factors that predict transitioning from resilient to symptomatic classes. Our aim is to (1) identify longitudinal classes (i.e., subgroups of individuals) based on indicators of PTSD, AD, and well-being in response to the pandemic and (2) examine predictors of transitioning between these subgroups.

**Methods and analysis:** We will conduct a three-wave longitudinal online survey-study of *n* ≥ 2000 adults from the general Dutch population. The first measurement occasion takes place six months after the start of the pandemic, followed by two follow-up measurements with six months intervals. Latent transition analysis will be used for data-analysis.

**Ethics and dissemination:** Ethical approval has been obtained from four Dutch universities. Longitudinal study designs are vital to monitor mental health (and predictors thereof) in the pandemic to develop preventive and curative mental health interventions. This study is carried out by researchers who are board members of the Dutch Society for Traumatic Stress Studies and is part of a pan-European study (initiated by the European Society for Traumatic Stress Studies) examining the impact of the pandemic in eleven countries. Results will be published in peer-reviewed journals and disseminated at conferences, via newsletters, and media-appearance among (psychotrauma-)professionals and the general public.

**Strengths and limitations of this study:**

- This is one of the first studies examining the mental health impact of the COVID-19 pandemic by focusing on negative and positive mental health in the general population.
- A longitudinal research design is used, which enable us to examine predictors of transitioning between mental health classes over three time points.
- A limitation of this study is that we used self-report measures, instead of clinical interviews, to assess mental health.

## Introduction

The impact of the COVID-19 pandemic on mental health has been profound. In recent reviews a variety of psychological problems have been identified across studies on COVID-19 pandemic-related mental health effects [1–4]. One of the most commonly reported mental health problems during the pandemic are disturbances in stress reactions, such as posttraumatic stress disorder (PTSD) symptoms and adjustment disorder (AD) [5–7]. For instance, a meta-analysis found a pooled prevalence of post-pandemic PTSD of 23% [8] and two studies from the UK and Poland reported AD prevalence rates during the COVID-19 pandemic of 16% and 49%, respectively [5,6]. These rates are higher than one-year prevalence rates of 4-5% PTSD [9,10] and point prevalence rate of 1% of AD [11] found in the general population before the pandemic.

PTSD and AD are both categorized as disorders specifically related to exposure to trauma and stress in the DSM-5 [12] and ICD-11 [13]. People with PTSD experience intense, disturbing thoughts and feelings related to a traumatic event that happened at least one month earlier. They may relive this event through flashbacks or nightmares and feel sadness, fear, anger, or feel detached from others [12]. AD has been characterized by “marked distress that is out of proportion to the severity or intensity of the stressor” [12]. This distress is represented by “preoccupation with the stressor or its consequences, including excessive worry, recurrent and distressing thoughts about the stressor, or constant rumination about its implications, as well as by failure to adapt to the stressor” [13]. Whereas the PTSD A-criterion requires the stressor to be related to death, threatened death, actual or threatened serious injury, or actual or threatened sexual violence, for AD the stressor may be associated with various psychosocial life-stressors such as divorce, illness or disability, socio-economic problems, or conflicts at home or work [12,13]. As both classification systems prescribe that AD typically does not last longer than six months and may not be explained better by another mental disorder, AD has been referred to as a subclinical or mild disorder compared with other psychiatric diagnoses [14]. Recent research suggests that AD may be an early marker for more severe disorders, such as PTSD [15].

So far, most of the research on the psychological impact of the COVID-19 pandemic has been focused on the presence of psychological disorders [1–4]. This approach does not capture the complete picture of mental health for at least two reasons. Firstly, focusing on mental illnesses only provides a limited perspective on mental health. As described by the World Health Organization (WHO), mental health is not only defined by the absence of psychopathology it “is defined as a state of well-being in which every individual realizes his or her own potential, can cope with the normal stresses of life, can work productively and fruitfully, and is able to make a contribution to her or his community” [16]. The dual-continua model states that well-being and symptoms of psychopathology are related, but separate continua [17]. Empirical work supports that increases in well-being are related to less psychopathology, however absence of psychopathology does not indicate a high level of well-being or vice versa [18–20]. Broadening our focus from mental illness to mental health by including psychopathology *and* well-being may therefore yield a more complete picture and better understanding of the psychological impact of the pandemic.

Secondly, mental disorders have often been investigated as either present or absent by reporting prevalence rates or by examining how people respond on average. These approaches ignore individual differences in psychological responses. An increasing body of research offers support that psychological responses are heterogeneous [21–25]. To illustrate this, there are 636,120 ways to have PTSD [26]. A statistical technique that has often been used to study heterogeneity in responses is latent class analysis (LCA). LCA categorizes individuals into classes based on similar response patterns. For instance, LCA has been used to show that mental illness and well-being are separate continua; classes were found of people showing high well-being and low psychopathology symptoms, whereas other classes were characterized by people with elevated symptoms while reporting high well-being [27].

While LCA might be helpful in identifying latent classes of people that differ in mental health indicators during the pandemic, this has, to the best of our knowledge, not been studied yet. We therefore aim to examine latent classes of AD, PTSD, and well-being in order to enhance our knowledge on the impact of the COVID-19 pandemic on mental health. As the pandemic is evolving, people’s mental health responses are changing as well. To capture this, longitudinal research is needed. Latent transition analysis (LTA) is a longitudinal extension of a LCA, which is helpful in capturing the fluctuating nature of symptom-profiles [24,25]. With LTA the likelihood of transitioning between classes over time is estimated, so for instance the likelihood of moving from the high well-being and low psychopathology symptoms class at the first measurement occasion to the high well-being and high psychopathology symptoms class at the second measurement occasion. Furthermore, predictors of transitions can be added to the LTA allowing identification of risk and protective factors for mental health. Examining predictors of transitions of mental health during the pandemic is relevant to identify people at risk for developing worsening of symptoms as well as identifying protective factors that enhance well-being. This knowledge is considered helpful for screening, prevention, and treatment of stress reactions and enhancement of well-being related to the global pandemic.

This study aims to examine longitudinal symptom profiles of PTSD, AD, and well-being in a general population sample in the Netherlands during the COVID-19 pandemic using data from three measurement occasions with a six month time interval. LCA and LTA will be used to examine longitudinal symptom profiles. Based on previous studies on individual differences in mental health [22,23,27] we expect to identify at least five classes that differ with respect to symptom-profiles. We expect that the modal response would consist of a subgroup representing complete mental health (i.e., individuals with low PTSD and AD symptomatology and moderate to high well-being) (class 1), based on prior research of symptom trajectories [22,23,27]. Furthermore, we expect to identify a class with severe symptomatology (high AD and PTSD symptoms) and low well-being (class 2) [21,27]. Considering findings on mental health prevalence from cross-sectional studies of COVID-19 pandemic responses [6], we also expect a class of individuals with difficulties adjusting (moderate to high AD symptoms) and moderate/low well-being, but who do not show traumatic responses (low PTSD symptoms; class 3), and a class for whom PTSD symptoms are at the forefront (high PTSD) and moderate/low well-being, but who experience moderate/low adjustment difficulties (class 4) [28,29]. Lastly, in line with the dual continua model, we expect a class that shows elevated AD and PTSD symptomatology while maintaining moderate to high well-being levels (class 5) [17,27].

Transitions between classes will be explored by including the following variables related to the participants’ (i) socio-demographics, (ii) profession, and (iii) health, based on prior research examining predictors of distress after adversity [30–34]. Regarding the socio-demographic variables, we expect individuals who are female, belong to an ethnic minority, and are exposed to childhood trauma are more likely to belong/transition to classes with more distress and low well-being. Furthermore, with respect to profession, we expect students and people in a healthcare profession to belong/transition to classes with pervasive distress. In addition, we expect that individuals with a history of mental health issues and poor self-reported health are at greater odds to belong to classes with poor mental health. We also expect individuals who report more exposure to pandemic-related stressors (i.e., being infected with COVID-19, experiencing a death of a loved one during the pandemic, loss of income due to the pandemic, or loss of social network) to be at risk for belonging/transitioning from asymptomatic to symptomatic and from high to low well-being [35].

## Methods and Analysis

### Study design

This is a longitudinal online cohort study, called CONNECT, carried out among participants from the Dutch general population. This study is part of a pan-European research collaboration under co-ordination of the European Society for Traumatic Stress Studies (ESTSS) conducted in eleven countries (for more details, see Lotzin et al., 2020) [36]. Participants will be assessed at three time points (i.e., T1, T2, T3), six months apart. Baseline assessment took place from July 16 to November 16, 2020.

### Eligibility

Participants need to be at least 18 years of age, resident in the Netherlands at the time of study participation, and able to read Dutch or English. No exclusion criteria will be applied. Participants will be informed about confidentiality and will be asked to provide informed consent before filling out the survey.

### Sample size

In line with the pan-European research study, the sample size will be set to *n*=2000 (i.e., *n* = 2000 participants in countries with more than 15 million inhabitants). We expect a 65% response rate per consecutive wave (T2: *n* = 1300, T3: *n* = 845).

### Recruitment and procedure

Participants will be recruited via social media platforms (e.g., LinkedIn, Facebook, Twitter, Instagram, WhatsApp), social networks of the authors, and mental health care clinics and universities whom the authors are affiliated with. Participants will be able to take part in a raffle to win a voucher at every assessment time point (chance of winning 1:100, 25€). First year students from Utrecht University, Leiden University, the University of Groningen, and Radboud University Nijmegen can participate in exchange for course credits. A marketing agency will be used to recruit 300 individuals from a volunteer panel of the general population who represent specific demographic subgroups that are commonly more difficult to include in scientific research: men, people with low to middle socioeconomic backgrounds, and those between 40 and 60 years of age. Eligible participants are provided with a weblink to the survey consisting of questionnaires and asked to complete it, which takes approximately 25 minutes.

### Measures

Detailed information on instruments used in the CONNECT Study can be found at Open Science Framework (https://osf.io/qeba5/). Below we describe the measures that will be used in the current study.

### Mental health indicators

PTSD symptoms will be assessed with the Primary Care PTSD screen for DSM-5 (PC-PTSD-5) [37], a 5-item, dichotomous (0 = no, 1 = yes) screening measure assessing symptoms (e.g., “Been constantly on guard, watchful, or easily startled?”) experienced over the past month. The PC-PTSD-5 showed good diagnostic accuracy in a sample of 398 US veterans in primary care [38].

AD symptoms will be assessed with the Adjustment Disorder – New Module 8 Questionnaire (ADNM-8) [39], an 8-item self-report measure. Items (e.g., “My thoughts often revolve around anything related to the stressful situation“) are rated on a 4-point Likert scale ranging from 1 (never) to 4 (often). Previous research indicated sound construct validity and good internal validity across help-seeking individuals with symptoms of AD [39].

Well-being is assessed with the WHO-Five Well-being Index, a derivative of the 28-item and 10-item WHO Well-Being Questionnaires [40]. The WHO-Five measures well-being over the past two weeks on a six-point Likert scale from 0 (at no time) to 5 (all the time). A sample question is “I have felt cheerful and in good spirits”. The WHO-5 has shown to be validated in a number of studies in various languages [41].

### Predictors of class-membership

With respect to socio-demographic characteristics, we will assess participants’ gender and ethnicity (categorized as Dutch vs. non-Dutch). Childhood trauma will be assessed with the Childhood Experience Questionnaire (ACE) [42], relating to emotional, physical, and sexual abuse, and emotional and physical neglect. Respondents rate whether the respective childhood trauma was experienced before the age of 19 (no or yes). The ACE has sound construct validity and good internal validity across samples, including clinical and nonclinical samples, and demonstrates convergent validity with the Childhood Trauma Questionnaire [43].

Related to the participants’ profession, we will ask “What is your current situation regarding education or employment?”. We will categorize this as student vs. non-student. In case people are employed we will ask in what field they work in (categorized as healthcare profession vs. other).

Questions regarding health will contain history of mental health issues (i.e., “Have you ever been diagnosed with a psychiatric disorder (such as a depressive or anxiety disorder?” yes or no) and current self-rated health (How would you describe your current health status?) categorized as satisfactory/good vs. poor/very poor.

The following stressors related to COVID-19 will be assessed with a set of items developed for the ESTSS COVID-19 Study [36]: being infected with COVID-19 (yes or no), experiencing a death of a loved one during the pandemic (yes or no), loss of income due to the pandemic (yes or no), and perceived burden of loss of social network (categorized as not at all/a bit vs. somewhat/severe).

### Data Analysis

Latent transition analysis (LTA) will be used for analyzing the data. The LTA will be performed in four consecutive steps. In the first step, latent class analyses (LCAs) will be conducted for each measurement occasion separately. In line with earlier studies using LTA and LCA [24,25,44,45] non-binary indicators (i.e., symptoms of adjustment disorder symptoms and well-being) will be dichotomized, such that ‘symptom absence’ is represented by scores of 0 for PTSD, 1 or 2 for adjustment disorder, and 0-2 for well-being and ‘symptom presence’ is characterized by scores of 1 on PTSD, 3 or 4 adjustment disorder, and 3-5 for well-being.

Latent class models up to eight classes will be run. The model with the best fit will be selected. Better model fit is indicated by: i) lower values for the Bayesian Information Criterion (BIC) and Akaike Information Criterion (AIC), ii) higher entropy R^2^ values, and iii) a significant *p*-value (<.05) of the Vuong–Lo–Mendell–Rubin test (VLMRt), Lo-Mendell-Rubin-Likelihood Ratio Test (LMR-LRt), and the Bootstrap-Likelihood Ratio Test (BLRt). We will also take the parsimony and interpretability of the models into account.

In the second step, we will examine measurement invariance between the classes at the separate measurement occasions. This will be tested to examine whether the number of the classes and symptom profiles of the classes are similar across measurement occasions [46]. A non-significant log-likelihood difference test will be used as an indicator for measurement invariance.

In the third step, we will examine transition probabilities by regression class-membership of one measurement occasion on the class-membership of the preceding measurement occasion. Transition probabilities are the probability of people staying in the same class over time or the probability of people transitioning from one class to another class at a subsequent time point.

In the fourth and final step, predictors will be added to the LTA using multi-nominal logistic regression analyses. In doing so, we can predict the likelihood of moving out of a class to another class at a subsequent time point compared with staying in the same class over time. Multiple imputations will be used to handle missing data on predictors [47].

## Data Availability

With consent from the participants, data will be deposited, stored, and shared at a secure data management service from Utrecht University.

https://osf.io/bxg68/

## Ethics and Dissemination

The study has been approved by the ethics committee of Utrecht University (20-360; T. Mooren), Leiden University (2020-09-10; J. Mouthaan-V1-2619), the University of Groningen (PSY-1920-S-0517; L. Lenferink), and Radboud University Nijmegen (ECSW-2020-127; M. Eidhof). All researchers involved in this study, except for AMF and SS, are board members of the Dutch Society for Traumatic Stress Studies [in Dutch: de Nederlandstalige Vereniging voor Psychotrauma]. This study is part of a pan-European research collaboration initiated by the European Society for Traumatic Stress Studies (ESTSS) including eleven countries (for more details Lotzin et al., 2020) [36]. The research findings will be published in a peer-reviewed, open access journal article and disseminated among researchers, clinicians, members of our society, and policy makers at conference talks, via news updates on the website of the Dutch Society for Traumatic Stress Studies (www.ntvp.nl), and media-appearances. With consent from the participants, data will be deposited, stored, and shared at a secure data management service from Utrecht University.

## Patient and Public involvement statement

Potential participants or the public were not involved in the design, or conduct, or reporting, or dissemination plans of our research.

## Authors’ contributions

Lonneke I.M. Lenferink conceptualized the study and together with J. Mouthaan drafted the first version of the manuscript. Prof. Trudy Mooren is principal investigator of this project. All other authors critically read and revised the manuscript.

## References

1 Clemente-Suárez VJ, Dalamitros AA, Beltran-Velasco AI, et al. Social and Psychophysiological Consequences of the COVID-19 Pandemic: An Extensive Literature Review. Front Psychol 2020;11:580225. doi:10.3389/fpsyg.2020.580225

2 Dubey S, Biswas P, Ghosh R, et al. Psychosocial impact of COVID-19. Diabetes Metab Syndr Clin Res Rev 2020;14:779–88. doi:10.1016/j.dsx.2020.05.035

3 Prati G, Mancini AD. The psychological impact of COVID-19 pandemic lockdowns: A review and meta-analysis of longitudinal studies and natural experiments. Psychol Med 2021;51:201–11. doi:10.1017/S0033291721000015

4 Vindegaard N, Benros ME. COVID-19 pandemic and mental health consequences: Systematic review of the current evidence. Brain Behav Immun 2020;89:531–42. doi:10.1016/j.bbi.2020.05.048

5 Ben-Ezra M, Hou WK, Goodwin R. Investigating the relationship between COVID-19-related and distress and ICD-11 adjustment disorder: two cross-sectional studies. BJPsych Open 2021;7. doi:10.1192/bjo.2020.158

6 Dragan M, Grajewski P, Shevlin M. Adjustment disorder, traumatic stress, depression and anxiety in Poland during an early phase of the COVID-19 pandemic. Eur J Psychotraumatol 2021;12. doi:10.1080/20008198.2020.1860356

7 Makhashvili N, Javakhishvili JD, Sturua L, et al. The influence of concern about COVID-19 on mental health in the Republic of Georgia: a cross-sectional study. Global Health 2020;16:1–10. doi:10.1186/s12992-020-00641-9

8 Yuan K, Gong YM, Liu L, et al. Prevalence of posttraumatic stress disorder after infectious disease pandemics in the twenty-first century, including COVID-19: a meta-analysis and systematic review. Mol Psychiatry 2021;:1–17. doi:10.1038/s41380-021-01036-x

9 Goldstein RB, Smith SM, Chou SP, et al. The epidemiology of DSM-5 posttraumatic stress disorder in the United States: results from the National Epidemiologic Survey on Alcohol and Related Conditions-III. Soc Psychiatry Psychiatr Epidemiol 2016;51:1137–48. doi:10.1007/s00127-016-1208-5

10 Kessler RC, Wai TC, Demler O, et al. Prevalence, severity, and comorbidity of 12-month DSM-IV disorders in the National Comorbidity Survey Replication. Arch Gen Psychiatry 2005;62:617–27. doi:10.1001/archpsyc.62.6.617

11 Maercker A, Forstmeier S, Pielmaier L, et al. Adjustment disorders: prevalence in a representative nationwide survey in Germany. Soc Psychiatry Psychiatr Epidemiol 2012 4711 2012;47:1745–52. doi:10.1007/S00127-012-0493-X

12 American Psychiatric Association. Diagnostic and statistical manual of mental disorders (5th ed.). Arlington, VA: American Psychiatric Association. 2013. doi:https://doi.org/10.1176/appi.books.9780890425596

13 World Health Organization. International statistical classification of diseases and related health problems (11th ed.). 2020. https://icd.who.int/

14 O’Donnell ML, Alkemade N, Creamer M, et al. A longitudinal study of adjustment disorder after trauma exposure. Am J Psychiatry 2016;173:1231–8. doi:10.1176/appi.ajp.2016.16010071

15 O’Donnell ML, Agathos JA, Metcalf O, et al. Adjustment disorder: Current developments and future directions. Int J Environ Res Public Health 2019;16. doi:10.3390/ijerph16142537

16 World Health Organization. WHO urges more investments, services for mental health. WHO. 2012.

17 Keyes CLM. Mental illness and/or mental health? Investigating axioms of the complete state model of health. J Consult Clin Psychol 2005;73:539–48. doi:10.1037/0022-006X.73.3.539

18 Westerhof G. The dual continua model of mental health and illness: Theory, findings, and applications in psychogerontology. In: Handbook of Gerontology Research Methods: Understanding successful aging. Taylor & Francis 2016. 79–94. doi:10.4324/9781315771533

19 Westerhof GJ, Keyes CLM. Mental Illness and Mental Health: The Two Continua Model Across the Lifespan. J Adult Dev 2009 172 2009;17:110–9. doi:10.1007/S10804-009-9082-Y

20 de Vos JA, Radstaak M, Bohlmeijer ET, et al. Having an eating disorder and still being able to flourish? Examination of pathological symptoms and well-being as two continua of mental health in a clinical sample. Front Psychol 2018;9:2145. doi:10.3389/fpsyg.2018.02145

21 Breslau N, Reboussin BA, Anthony JC, et al. The structure of posttraumatic stress disorder: Latent class analysis in 2 community samples. Arch Gen Psychiatry 2005;62:1343–51. doi:10.1001/archpsyc.62.12.1343

22 Galatzer-Levy IR, Huang SH, Bonanno GA. Trajectories of resilience and dysfunction following potential trauma: A review and statistical evaluation. Clin Psychol Rev 2018;63:41–55. doi:10.1016/j.cpr.2018.05.008

23 Pierce M, McManus S, Hope H, et al. Mental health responses to the COVID-19 pandemic: a latent class trajectory analysis using longitudinal UK data. The Lancet Psychiatry 2021;0. doi:10.1016/S2215-0366(21)00151-6

24 Forbes D, Alkemade N, Nickerson A, et al. Prediction of late-onset psychiatric disorder in survivors of severe injury: Findings of a latent transition analysis. J Clin Psychiatry 2016;77:807–12. doi:10.4088/JCP.15m09854

25 Tay AK, Jayasuriya R, Jayasuriya D, et al. Twelve-month trajectories of depressive and anxiety symptoms and associations with traumatic exposure and ongoing adversities: a latent trajectory analysis of a community cohort exposed to severe conflict in Sri Lanka. Transl Psychiatry 2017;7:e1200. doi:10.1038/tp.2017.166

26 Galatzer-Levy IR, Bryant RA. 636,120 Ways to Have Posttraumatic Stress Disorder. Perspect Psychol Sci 2013;8:651–62. doi:10.1177/1745691613504115

27 Petersen KJ, Humphrey N, Qualter P. Latent Class Analysis of Mental Health in Middle Childhood: Evidence for the Dual-Factor Model. School Ment Health 2020;12:786–800. doi:10.1007/s12310-020-09384-9

28 Karatzias T, Shevlin M, Hyland P, et al. ICD-11 posttraumatic stress disorder, complex PTSD and adjustment disorder: the importance of stressors and traumatic life events. Anxiety, Stress Coping 2021;34:191–202. doi:10.1080/10615806.2020.1803006

29 Maercker A, Forstmeier S, Enzler A, et al. Adjustment disorders, posttraumatic stress disorder, and depressive disorders in old age: findings from a community survey. Compr Psychiatry 2008;49:113–20. doi:10.1016/j.comppsych.2007.07.002

30 Fruehwirth JC, Biswas S, Perreira KM. The Covid-19 pandemic and mental health of first-year college students: Examining the effect of Covid-19 stressors using longitudinal data. PLoS One 2021;16:e0247999. doi:10.1371/journal.pone.0247999

31 Killikelly C, Lenferink LIM, Xie H, et al. Rapid Systematic Review of Psychological Symptoms in Health Care Workers COVID-19. J Loss Trauma Published Online First: 2021. doi:10.1080/15325024.2020.1864115

32 Olff M. Sex and gender differences in post-traumatic stress disorder: an update. Eur J Psychotraumatol 2017;8:1351204. doi:10.1080/20008198.2017.1351204

33 Ozer EJ, Best SR, Lipsey TL, et al. Predictors of posttraumatic stress disorder and symptoms in adults: A meta-analysis. Psychol Bull 2003;129:52–73. doi:10.1037/0033-2909.129.1.52

34 Ryder AL, Azcarate PM, Cohen BE. PTSD and Physical Health. Curr Psychiatry Rep 2018;20:1–8. doi:10.1007/s11920-018-0977-9

35 McGinty EE, Presskreischer R, Anderson KE, et al. Psychological distress and covid-19– related stressors reported in a longitudinal cohort of US adults in April and July 2020. JAMA - J Am Med Assoc 2020;324:2555–7. doi:10.1001/jama.2020.21231

36 Lotzin A, Acquarini E, Ajdukovic D, et al. Stressors, coping and symptoms of adjustment disorder in the course of the COVID-19 pandemic – study protocol of the European Society for Traumatic Stress Studies (ESTSS) pan-European study. Eur J Psychotraumatol 2020;11:1780832. doi:10.1080/20008198.2020.1780832

37 Prins A, Bovin MJ, Kimerling R, et al. Primary Care PTSD Screen for DSM-5 (PC-PTSD-5). Natl Cent PTSD 2015;5:1–3.https://www.ptsd.va.gov/professional/%0Ahttps://www.ptsd.va.gov/professional/assessment/screens/pc-ptsd.asp (accessed 22 May 2021).

38 Prins A, Bovin MJ, Smolenski DJ, et al. The Primary Care PTSD Screen for DSM-5 (PC-PTSD-5): Development and Evaluation Within a Veteran Primary Care Sample. J Gen Intern Med 2016;31:1206–11. doi:10.1007/s11606-016-3703-5

39 Kazlauskas E, Gegieckaite G, Eimontas J, et al. A Brief Measure of the International Classification of Diseases-11 Adjustment Disorder: Investigation of Psychometric Properties in an Adult Help-Seeking Sample. Psychopathology 2018;51:10–5. doi:10.1159/000484415

40 Bech P. Measuring the Dimension of Psychological General Well-Being by the WHO-5. Qual Life Newsl 2004;32:15–6.

41 Topp CW, Østergaard SD, Søndergaard S, et al. The WHO-5 well-being index: A systematic review of the literature. Psychother Psychosom 2015;84:167–76. doi:10.1159/000376585

42 Felitti VJ, Anda RF, Nordenberg D, et al. Relationship of childhood abuse and household dysfunction to many of the leading causes of death in adults: The adverse childhood experiences (ACE) study. Am J Prev Med 1998;14:245–58. doi:10.1016/S0749-3797(98)00017-8

43 Schmidt MR, Narayan AJ, Atzl VM, et al. Childhood Maltreatment on the Adverse Childhood Experiences (ACEs) Scale versus the Childhood Trauma Questionnaire (CTQ) in a Perinatal Sample. J Aggress Maltreatment Trauma 2020;29:38–56. doi:10.1080/10926771.2018.1524806

44 Lenferink, L.I.M., Liddell, B.J., Byrow, Y., O’Donnell, M., Bryant, R.A., Mau, V., McMahon, T., Benson, G., & Nickerson A. Course and predictors of posttraumatic stress and depression longitudinal symptom profiles in refugees: A latent transition model. Submitted

45 Ulbricht CM, Chrysanthopoulou SA, Levin L, et al. The use of latent class analysis for identifying subtypes of depression: A systematic review. Psychiatry Res 2018;266:228–46. doi:10.1016/j.psychres.2018.03.003

46 Nylund KL, Muthén B, Nishina A, et al. Stability and instability of peer victimization during middle school: Using latent transition analysis with covariates, distal outcomes, and modeling extensions. Unpublished manuscript. 2006;:1–50.https://www.statmodel.com/download/LTA_DP_FINAL.pdf (accessed 24 May 2021).

47 Little TD. Longitudinal Structural Equation Modeling. 2013.

